# Predictors of SARS-CoV-2 infection following high-risk exposure

**DOI:** 10.1101/2021.10.20.21265295

**Authors:** Kristin L. Andrejko, Jake Pry, Jennifer F. Myers, John Openshaw, James Watt, Nozomi Birkett, Jennifer L. DeGuzman, Sophia S. Li, Camilla M. Barbaduomo, Anna T. Fang, Vivian H. Tran, Mahsa H. Javadi, Paulina M. Frost, Zheng N. Dong, Seema Jain, Joseph A. Lewnard, on behalf of the California COVID-19 Case-Control Study Team

## Abstract

**Background:** Non-pharmaceutical interventions (NPIs) are recommended for COVID-19 mitigation. However, the effectiveness of NPIs in preventing SARS-CoV-2 transmission remains poorly quantified.

**Methods:** We conducted a test-negative design case-control study enrolling cases (testing positive for SARS-CoV-2) and controls (testing negative) with molecular SARS-CoV-2 diagnostic test results reported to California Department of Public Health between 24 February-26 September, 2021. We used conditional logistic regression to assess predictors of case status among participants who reported contact with an individual known or suspected to have been infected with SARS-CoV-2 (“high-risk exposure”) within ≤14 days of testing.

**Results:** 643 of 1280 cases (50.2%) and 204 of 1263 controls (16.2%) reported high-risk exposures ≤14 days before testing. Adjusted odds of case status were 2.94-fold (95% confidence interval: 1.66-5.25) higher when high-risk exposures occurred with household members (vs. other contacts), 2.06-fold (1.03-4.21) higher when exposures occurred indoors (vs. not indoors), and 2.58-fold (1.50-4.49) higher when exposures lasted ≥3 hours (vs. shorter durations) among unvaccinated and partially-vaccinated individuals; excess risk associated with such exposures was mitigated among fully-vaccinated individuals. Mask usage by participants or their contacts during high-risk exposures reduced adjusted odds of case status by 48% (8-72%). Adjusted odds of case status were 68% (32-84%) and 77% (59-87%) lower for partially- and fully-vaccinated participants, respectively, than for unvaccinated participants. Benefits of mask usage were greatest when exposures lasted ≥3 hours, occurred indoors, or involved non-household contacts.

**Conclusions:** NPIs reduced the likelihood of SARS-CoV-2 infection following high-risk exposure. Vaccine effectiveness was substantial for partially and fully vaccinated persons.

**KEY POINTS:**

- SARS-CoV-2 infection risk was greatest for unvaccinated participants when exposures to known or suspected cases occurred indoors or lasted ≥3 hours.
- Face mask usage when participants were exposed to a known or suspect case reduced odds of infection by 48%.

## INTRODUCTION

Strategies aimed at reducing risk of SARS-CoV-2 transmission during contact between infectious and susceptible individuals have been critical to mitigating the COVID-19 pandemic. While vaccines effectively reduce individual risk of infection and severe disease [1–3], non-pharmaceutical interventions (NPIs) continue to be recommended in various circumstances; these include within populations ineligible for vaccination, in settings where vaccines remain inaccessible or under-utilized, and in response to SARS-CoV-2 variants with increased transmissibility. Efforts to prevent transmission include social distancing and avoiding direct physical contact with non-household members [4]; interacting with non-household members outdoors [5]; and use of face coverings to filter virus-containing droplets and aerosols [6,7].

However, evidence demonstrating how well various NPIs mitigate transmission risk remains limited [8,9]. Understanding of exposures mediating SARS-CoV-2 transmission stems largely from anecdotal reports with unknown generalizability [10]. Additionally, many assessments of the effectiveness of NPIs have been ecological studies comparing COVID-19 incidence before and after implementation of multiple interventions [11–13], making it difficult to distinguish effects of each strategy [14]. While numerous studies demonstrate that face masks limit the quantity of virus shed into the environment by infectious individuals [15,16], few have assessed real-world effectiveness of face masks in preventing SARS-CoV-2 infection [6]. Improved understanding of aspects of social contact that exacerbate or reduce risk of SARS-CoV-2 transmission are needed to guide intervention prioritization [17,18].

To mitigate transmission of SARS-CoV-2, California mandated social distancing and wearing of facial coverings in spring 2020, and implemented a tiered system for closure and reopening of public places based on community-level measures of SARS-CoV-2 transmission and hospital utilization [19]. Statewide social distancing and mask mandates among vaccinated people in most public places and the tiered system were relaxed on 15 June, 2021, when roughly 57% of eligible Californians were considered fully vaccinated [19,20]. However, amid rising incidence of COVID-19 and increases in hospitalizations following emergence of the Delta (B.1.617.2) variant [21,22], measures encouraging or requiring face masks in certain indoor settings regardless of vaccination status were reinstated [23]. We initiated a retrospective case-control study to understand risk factors for SARS-CoV-2 infection in California and inform public health strategies [1]. Here, we report predictors of SARS-CoV-2 infection among participants who reported high-risk exposures, defined as social contact with an individual known or suspected to have been infected with SARS-CoV-2 within two weeks preceding participants’ SARS-CoV-2 tests.

## METHODS

### Design

California residents with confirmatory, molecular SARS-CoV-2 diagnostic test results reported to the California Department of Public Health (CDPH) between 24 February, 2021 and 26 September, 2021 with a recorded phone number were eligible for inclusion. Each day, interviewers called participants selected at random from all individuals with test results reported in the preceding 48 hours. Cases were persons with a positive molecular SARS-CoV-2 test result while controls were persons with a negative result. We enrolled cases equally across nine regions of the state (**Table S1**). For each enrolled case, interviewers attempted to enroll one control matched to the case by age group, sex, region, and week of SARS-CoV-2 test from a list of ≥30 randomly selected controls meeting these criteria. Participants were eligible to enroll if they provided informed consent in English or Spanish, and had not received a previous diagnosis of COVID-19 or positive test result for SARS-CoV-2 infection (molecular, antigen, or serological test). Additional sampling and enrollment details have been described elsewhere [1].

The study protocol was approved as public health surveillance by the State of California Health and Human Services Agency Committee for the Protection of Human Subjects.

### Exposures

Interviewers administered a standardized phone-based questionnaire to assess exposures (**Text S1**). This analysis included participants who reported they were potentially exposed to SARS-CoV-2 within 14 days prior to their test through social contact with an individual known or suspected by the participant to have been infected with SARS-CoV-2 at the time of their interaction (“high-risk exposure”). Participants were asked to specify if they were aware that one or more of these individuals had been a confirmed case, based on receipt of a positive diagnostic test result for SARS-CoV-2 infection. As a confirmatory check, we repeated the analyses described below using only the subset of participants who reported contact with a confirmed case.

Among participants reporting high-risk exposure, interviewers documented exposure attributes including setting (any indoor exposure versus outdoor exposure only); duration (whether contact lasted ≥3 hours); whether the participant and the contact had any physical contact; whether the contact was a household member; and use of face coverings by the participant and the contact during the interaction(s).

Additionally, all study participants were asked to indicate their reasons for seeking SARS-CoV-2 testing, including any symptoms experienced in the 14 days preceding their test. Interviews also addressed participants’ self-reported history of visiting other locations, including restaurants, bars, coffee shops, retail shops, public gyms, salons, movie theaters, or worship services; participating in social gatherings; and using ride share services, public transportation, or air travel. Interviewers recorded the COVID-19 vaccination status of participants, including the manufacturer and dates of all doses received.

### Statistical analysis

Our primary analysis aimed to identify characteristics of high-risk exposure associated with SARS-CoV-2 infection among participants. We fit a conditional logistic regression model to estimate adjusted odds ratios (aORs) and accompanying 95% confidence intervals (CIs) of various exposure attributes, comparing cases with controls. These included exposure setting (any indoor exposure versus outdoor-only exposure), exposure duration (any exposure ≥3 hours versus <3 hours), whether the exposure involved a potentially infectious household member(s) (versus non-household contact(s) only), the nature of exposure (any physical contact versus no physical contact), and mask usage by the participant or their contact during the entire interaction (versus mask usage by neither party). Models included interaction terms between each contact attribute and the vaccination history of the participant at the time of their test to assess effect modification. We considered participants tested >14 days after receipt of two doses of BNT162b2 (Pfizer/BioNTech) or mRNA-1273 (Moderna) or one dose of JNJ-78436735 (Janssen Pharmaceutical Companies) to be fully vaccinated. Others reporting receipt of any COVID-19 vaccine doses before their test date were considered partially vaccinated.

To correct for differences in infection prevalence over time and across regions, independent of the specific exposures being analyzed, regression strata were defined by the reopening tier of participants’ county of residence at the time of testing, or, for the period after 15 June, 2021 (when the tiered reopening system was retired), by participants’ month of SARS-CoV-2 testing. We further controlled for potential confounders including demographic variables (age, sex, and region), and participants’ reported attendance at community settings (as listed above) which may have been associated with risk of SARS-CoV-2 exposure.

We also undertook secondary analyses estimating the aOR of mask usage among cases versus controls within specific high-risk exposure strata [24]. Consistent with our primary analyses, these included indoor and outdoor exposures, ≥3 hour and <3 hour exposures, exposures to potentially infected individuals who were and were not members of participants’ households, and exposures with and without physical contact. We further estimated the aOR of mask usage separately among fully-vaccinated and partially-vaccinated or unvaccinated participants. Conditional logistic regression models for these analyses followed the framework described above and included interaction terms between each exposure characteristic and mask usage by participants or their contacts.

Last, we tested the hypothesis that face mask usage during high-risk exposure reduces the severity of illness among SARS-CoV-2 infected individuals [25–27]. We restricted our analytic sample to cases testing positive for SARS-CoV-2. As measures of severity, we considered whether participants reported any COVID-19 symptoms, and whether they reported any type of consultation with a health care provider (e.g., virtual or outpatient appointment, emergency room attendance, or hospitalization) in conjunction with testing. We used logistic regression models controlling for the same variables listed above to estimate aORs of mask-wearing by the participant and their contact, comparing cases with symptoms to those without symptoms and cases who sought healthcare (beyond diagnostic testing) to those who did not. As these analyses were limited to individuals testing positive for SARS-CoV-2 infection, the conditional logistic regression framework used in the primary analyses to adjust for differences in infection prevalence across locations and time was unnecessary.

We conducted analyses in R software (version 3.6.1).

## RESULTS

### Enrollment and descriptive analyses

Between February 24 and September 26, 2021, we enrolled 2541 participants, including 1279 cases and 1262 controls. In total, 847 participants, including 643 cases (50% of 1279) and 204 controls (16% of 1262), reported high-risk exposure within 14 days before testing, including 694 (82% of 847) with confirmed and 153 (18% of 847) with suspected exposure (**Table 1; Table S2**). Most participants reported their high-risk exposure occurred within a household (55% of 847) or workplace (14% of 847) (**Table S3**). A majority of these participants (69%, 582/847) listed high-risk exposure as a motivation for testing; additionally, 280 (33% of 847) participants sought testing due to symptoms (**Table S4**).

**Table 1.** Descriptive attributes of participants reporting high-risk exposures.

|  |  | All participants<br>*n* (%)<br>*N*=847 | Cases<sup>1</sup><br>*n* (%)<br>*N*=643 | Controls<sup>2</sup><br>*n* (%)<br>*N*=204 |
| --- | --- | --- | --- | --- |
| Age | 0-6 | 30 (3.5) | 28 (4.4) | 2 (1.0) |
|  | 7-12 | 48 (5.7) | 37 (5.8) | 11 (5.4) |
|  | 13-17 | 53 (6.3) | 41 (6.4) | 12 (5.9) |
|  | 18-29 | 269 (31.8) | 206 (32.0) | 63 (30.9) |
|  | 30-49 | 291 (34.4) | 202 (31.4) | 89 (43.6) |
|  | 50-64 | 111 (13.1) | 91 (14.2) | 20 (9.8) |
|  | 65+ | 45 (5.3) | 38 (5.9) | 7 (3.4) |
| Sex | Male | 388 (45.8) | 295 (45.9) | 93 (45.6) |
|  | Female | 459 (54.2) | 348 (54.1) | 111 (54.4) |
| Household income | Under \$50,000 | 206 (24.3) | 159 (24.7) | 47 (23.0) |
| | \$50,000 to \$100,000 | 201 (23.7) | 160 (24.9) | 41 (20.1) |
| | \$100,000 to \$150,000 | 99 (11.7) | 63 (9.8) | 36 (17.6) |
| | Over \$150,000 | 103 (12.2) | 73 (11.4) | 30 (14.7) |
|  | Refuse | 146 (17.2) | 114 (17.7) | 32 (15.7) |
|  | Not sure | 92 (10.9) | 74 (11.5) | 18 (8.8) |
| Race/ ethnicity | Non-Hispanic White | 357 (43.9) | 272 (44.2) | 85 (43.1) |
|  | Non-Hispanic Black | 38 (4.7) | 32 (5.2) | 6 (3.0) |
|  | Hispanic (any race) | 230 (28.3) | 176 (28.6) | 54 (27.4) |
|  | Asian | 67 (8.2) | 51 (8.3) | 16 (8.1) |
|  | Native American | 17 (2.1) | 15 (2.4) | 2 (1.0) |
|  | Native Hawaiian | 5 (0.6) | 5 (0.8) | 0 (0.0) |
|  | More than 1 race | 99 (12.2) | 65 (10.6) | 34 (17.3) |
|  | Refuse | 34 (4.1) | 27 (4.2) | 7 (3.4) |
| Region of residence<sup>3</sup> | *Predominantly urban regions* |  |  |  |
|  | San Francisco Bay Area | 87 (10.3) | 73 (11.4) | 14 (6.9) |
|  | Greater Los Angeles Area | 89 (10.5) | 70 (10.9) | 19 (9.3) |
|  | Greater Sacramento Area | 107 (12.6) | 82 (12.8) | 25 (12.3) |
|  | San Diego and southern border | 80 (9.4) | 62 (9.6) | 18 (8.8) |
|  | *Predominantly rural regions* |  |  |  |
|  | Central Coast | 113 (13.3) | 79 (12.3) | 34 (16.7) |
|  | Northern Sacramento Valley | 88 (10.4) | 69 (10.7) | 19 (9.3) |
|  | San Joaquin Valley | 92 (10.9) | 65 (10.1) | 27 (13.2) |
|  | Northwestern California | 96 (11.3) | 68 (10.6) | 28 (13.7) |
|  | Sierras Region | 95 (11.2) | 75 (11.7) | 20 (9.8) |
| Vaccination status<sup>4</sup> | Unvaccinated | 591 (72.0) | 502 (79.1) | 89 (47.8) |
|  | Partially vaccinated | 72 (8.8) | 49 (7.7) | 23 (12.4) |
|  | Fully vaccinated | 158 (19.2) | 84 (13.2) | 74 (39.8) |
| County reopening tier<sup>5</sup> | Purple tier (most restrictive) | 201 (23.7) | 160 (24.9) | 41 (20.1) |
|  | Red tier | 203 (24.0) | 163 (25.3) | 40 (19.6) |
|  | Orange tier | 200 (23.6) | 165 (25.7) | 35 (17.2) |
|  | Yellow tier (least restrictive) | 23 (2.7) | 17 (2.6) | 6 (2.9) |
|  | After June 15<sup>th</sup> | 220 (26.0) | 138 (21.5) | 82 (40.2) |
| Symptoms experienced | No symptoms |  |  |  |
|  | At least one symptom | 336 (39.7) | 178 (27.7) | 158 (77.5) |

Recent high-risk exposure is defined as reported contact with an individual known or suspected to have been infected with SARS-CoV-2 at any time within the 14 days before participants were tested.
<sup>1</sup>Cases reporting high-risk exposure represent 50% of 1280 cases who enrolled in and successfully completed the study.
<sup>2</sup>Controls reporting high-risk exposure represent 16% of 1263 controls who enrolled in and successfully completed the study.
<sup>3</sup>We list counties grouped into each region in **Table S1**.
<sup>4</sup>We defined participants as fully vaccinated at the time of their test if ≥14 days had passed following receipt of a second dose of BNT162b2 or mRNA-1273 (72 cases, 67 controls) or a single dose of JNJ-78436735 (12 cases, 7 controls). Participants who had received at least one dose of any COVID-19 vaccine, but did not meet these criteria for fully-vaccinated status, were considered partially vaccinated (46 cases and *N*=20 controls who received BNT162b2 or mRNA-1273; 3 cases and 3 controls who received JNJ-78436735). Participants who had not received any COVID-19 vaccine doses were considered unvaccinated.
<sup>5</sup>The State of California implemented a tiered system of reopening to reduce risk of SARS-CoV-2 transmission in community settings. On June 15, 2021, California discontinued the tiered system, relaxed facial masking requirements in certain indoor settings, and allowed businesses to reopen without physical distancing restrictions.

Among the 847 participants reporting high-risk exposure, 743 (88%) reported contact occurring indoors, 613 (72%) reported contact lasting ≥3 consecutive hours, 492 (58%) reported physical contact with the individual known or suspected to have been infected, and 385 (46%) indicated their contact was a household member. Participants who reported interactions occurring indoors, lasting ≥3 hours, or involving physical contact were generally more likely to have been enrolled after June 15, or to have resided in counties within less-restrictive reopening tiers at the time of their test, than those who reported outdoor, shorter, or non-physical contact (**Table 2**).

**Table 2.**
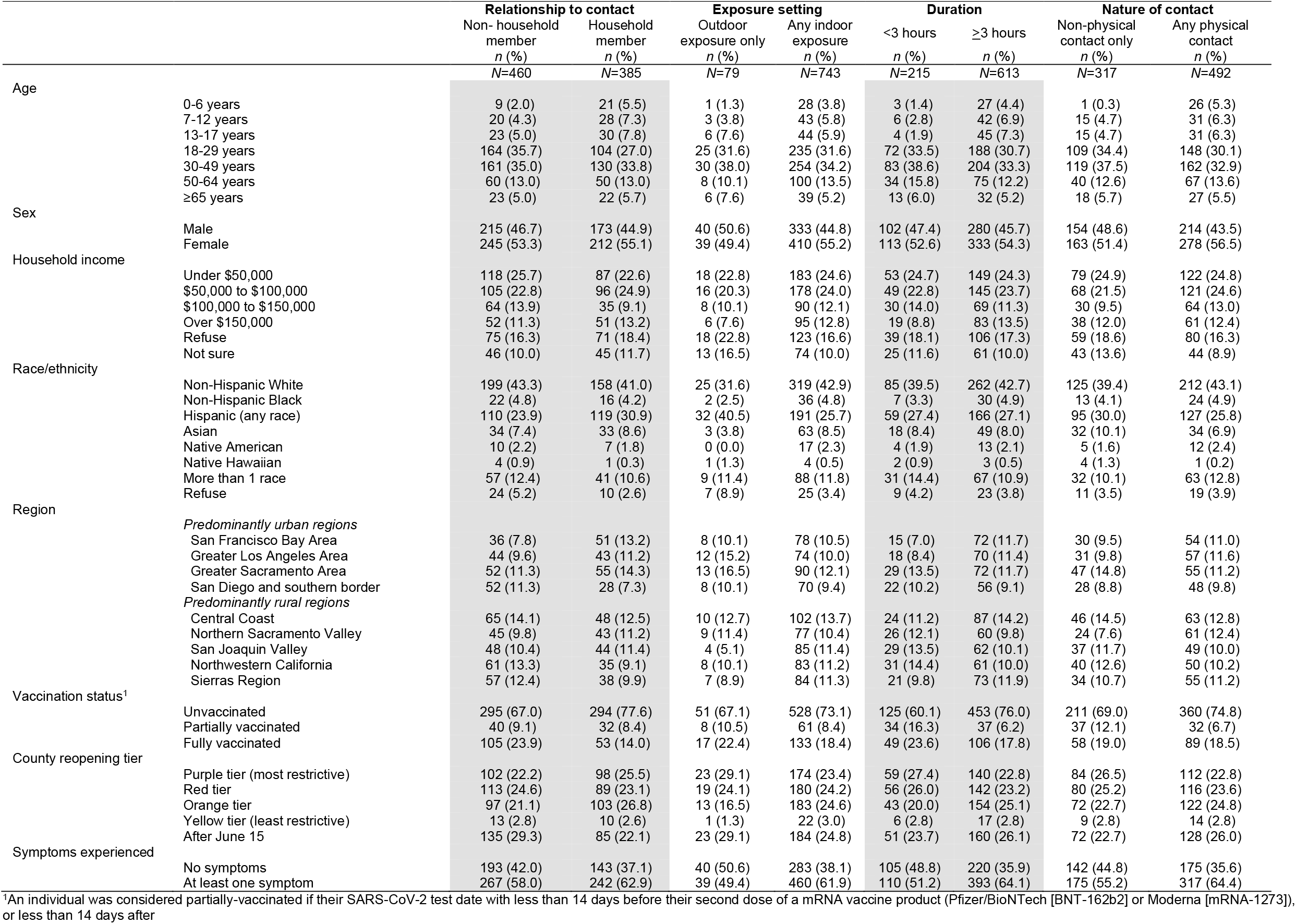
Attributes of participants reporting high-risk exposure with differing characteristics of contact.

The majority (82%, 694/847) of participants who had high-risk exposure reported both they and their contact did not wear a mask during the interaction (**Table 3**). Most participants were unvaccinated (70%, 591/847) at the time of testing; 9% (72/847) and 19% (158/847) were partially or fully vaccinated, respectively.

**Table 3.**
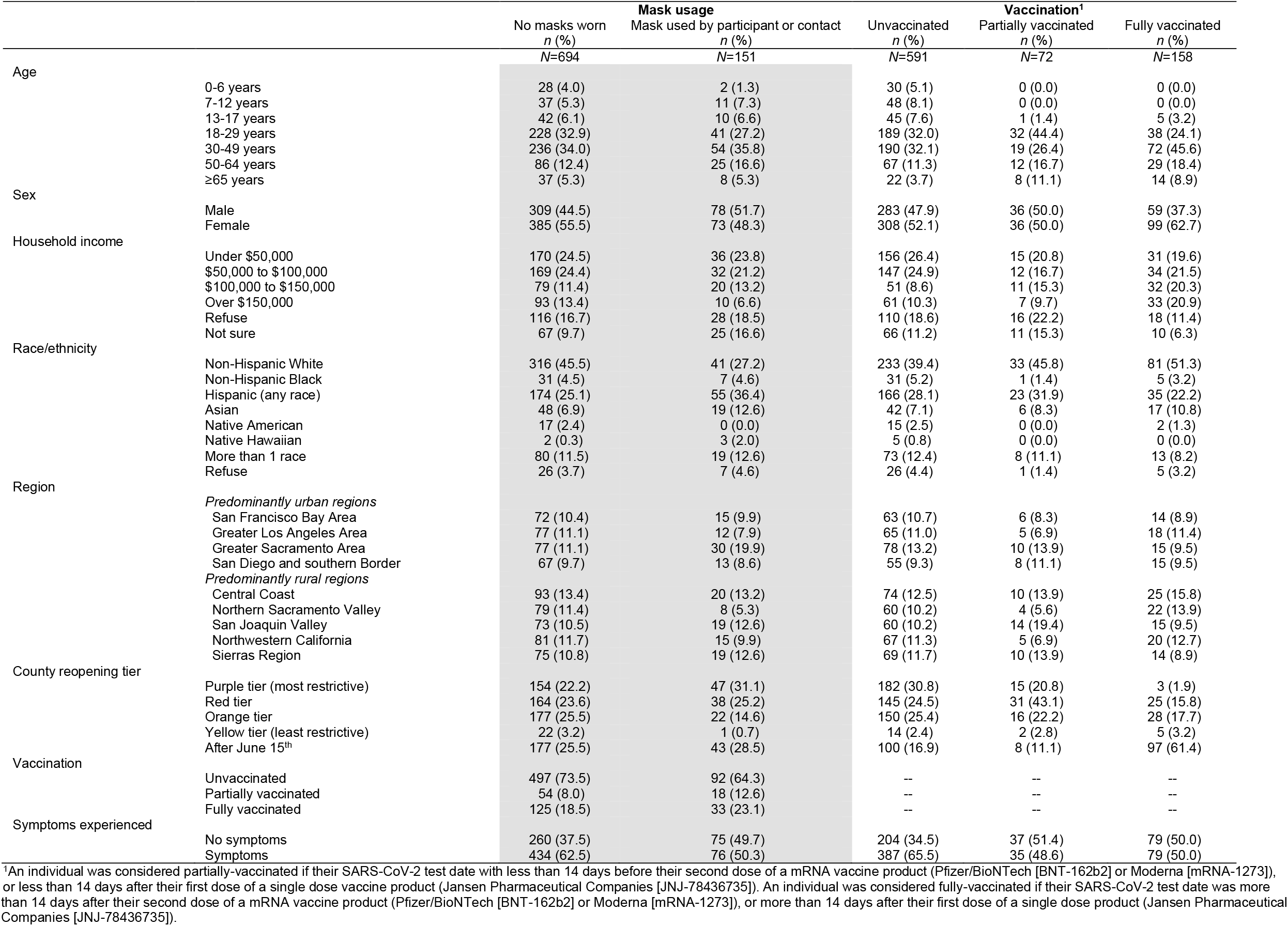
Distribution of exposures among respondents reporting differing types of recent contact with an individual known or suspected to have SARS-CoV-2 infection.

### Predictors of infection

Among unvaccinated or partially-vaccinated participants, cases were more likely to report high-risk exposures involving a potentially-infected household member, occurring indoors, lasting ≥3 hours, or where either they or their contact did not wear a face mask (**Figure 1**; **Table S5**). Estimated aORs of contact having occurred indoors, having lasted ≥3 hours, and having occurred with a household member were 2.06 (95% CI: 1.03-4.21), 2.58 (1.50-4.49), and 2.94 (1.66-5.25) fold higher among cases than controls, respectively. In contrast, we did not identify an association between case status and whether participants reported physical contact with the individual known or suspected to being infected. The association of each of these exposures with case status was mitigated among fully-vaccinated participants; within this sub-sample, the exposure associated with the greatest increase in adjusted odds of case status was contact with a household member (as compared to a non-household member; aOR=1.97 [0.88-4.55]). Estimated aORs were similar in models restricted to participants who specified that their contact was confirmed to have been infected with SARS-CoV-2 at the time of their interaction (**Table S6; Table S7**).

**Figure 1.**
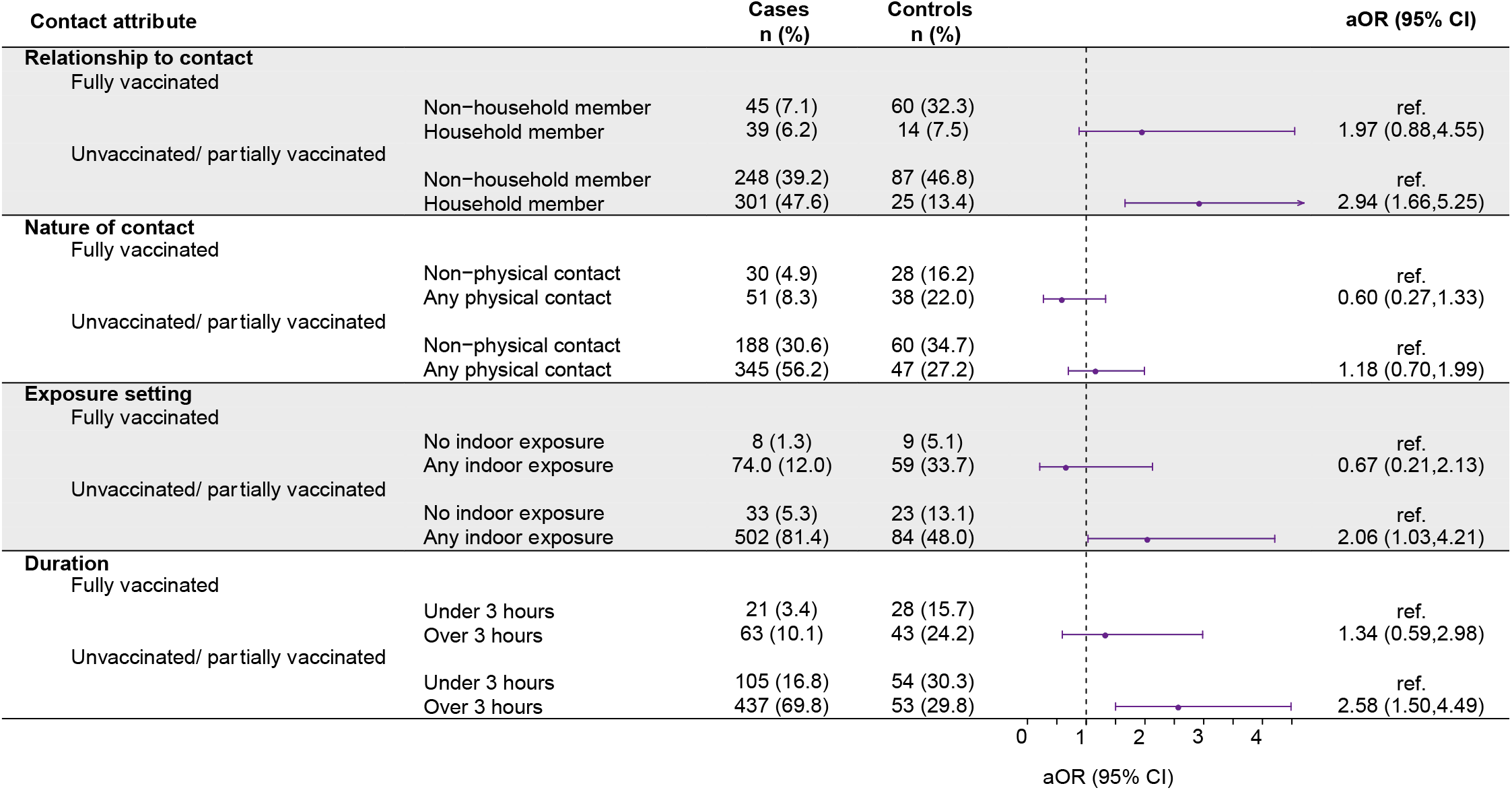
Predictors of infection following high-risk exposure. aOR: adjusted odds ratio, computed using conditional logistic regression models interacting vaccination status with each contact attribute, and adjusting for community exposures (listed in the main text), vaccination status (defined as fully vaccinated or unvaccinated/ incompletely vaccinated) of the participant and mask-wearing by the participant and their contact (as listed in **Figure 2**), and participants’ age, sex, and region of residence. Regression strata were defined for county reopening tiers and, for the period after June 15^th^, the month of SARS-CoV-2 test. Further regression parameter estimates are presented in **Table S4**. Counts for cases and controls differ from **Table 1** due to some participates indicating they did not know these details about their known or suspected contact, and missing data on vaccination status among cases (*N*=8) and controls(*N*=18). Due to occasional missing data, the denominators differ for the following counts: relationship of participant to contact among cases and controls (*N*=633, *N*=186), occurrence of physical contact among cases and controls (*N*=614, *N*=173), number of cases and controls reporting setting (*N*=617, *N*=175), number of cases and controls reporting duration of interaction (*N*=626, *N*=178).

Adjusted odds of mask usage by either participants or their contacts during high-risk interactions were 48% (9-71%) lower among cases than among controls (**Figure 2**). Estimated effect sizes did not differ appreciably according to whether masks were worn by participants, their contacts, or both, although analyses were underpowered to demonstrate significant effects within each of these strata or to make comparisons across them. Adjusted odds of having received a partial or full vaccination series were 68% (32-84%) and 77% (59-87%) lower among cases than among controls, respectively.

**Figure 2.**
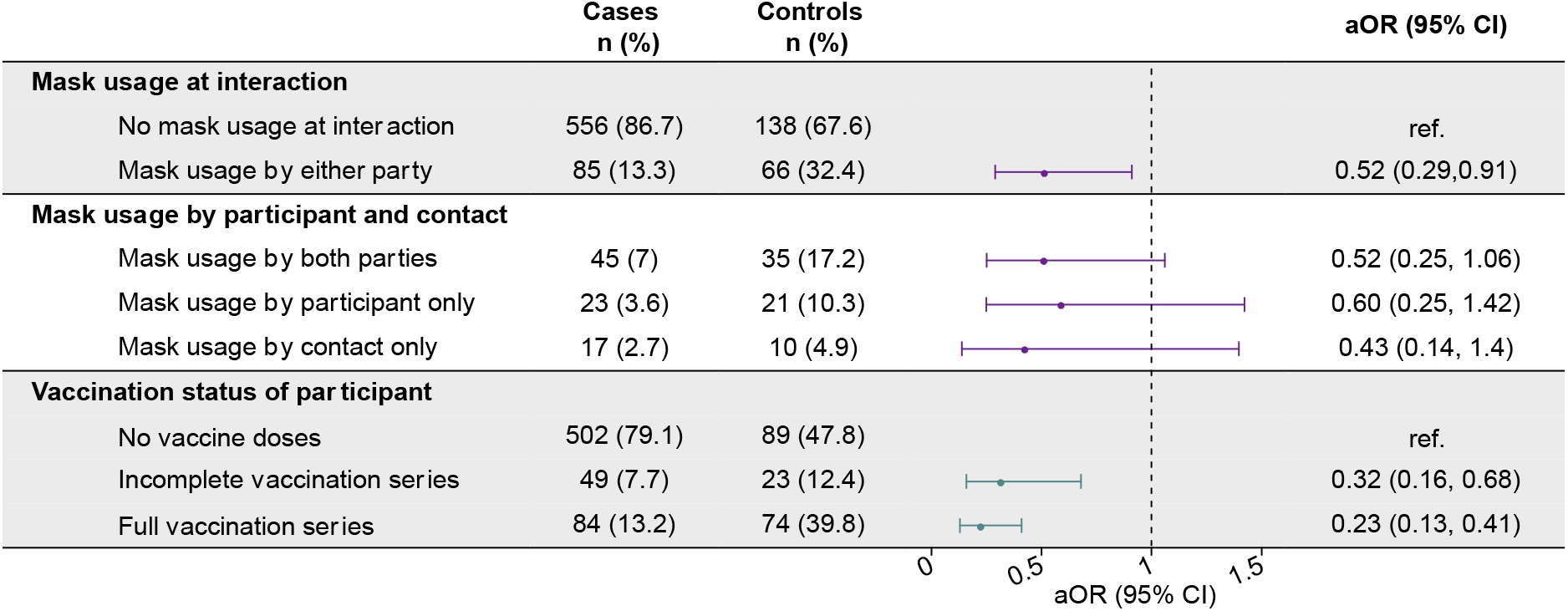
Protective effects of mask-wearing and vaccination in the context of high-risk exposure. aOR: adjusted odds ratio, computed using conditional logistic regression models adjusting for vaccination status, community exposures (listed in the main text), characteristics of high-risk contact (as listed in **Figure 1**), and participants’ age, sex, and region of residence. Regression strata were defined for county reopening tiers and week of SARS-CoV-2 test. Further regression parameter estimates are presented in **Table S4**. Due to occasional missing data, the denominators differ for the following counts: mask usage at interaction among cases and controls(*N*=641, *N*=204), vaccination status among cases and controls (*N*=635, *N*=186). An individual was considered fully vaccinated if their SARS-CoV-2 test date was more than 14 days after their second dose of a mRNA vaccine product (Pfizer/BioNTech [BNT-162b2] or Moderna [mRNA-1273]), or more than 14 days after their first dose of a single dose product (Jansen Pharmaceutical Companies [JNJ-78436735]). In sensitivity analyses limiting to those who received a mRNA vaccine product (excluding *N*=25 recipients of JNJ-78436735) the aOR (95% CI) for incompletely vaccinated and fully vaccinated individuals was 0.33 (0.15, 0.72) and 0.23 (0.12, 0.43), respectively.

Protective effects of mask usage by either participants or their contacts differed according to several characteristics of exposure events. Adjusted odds of mask usage were 53% (11-75%) lower among cases than among controls reporting indoor exposures, whereas statistically-significant effects of mask usage were not apparent among participants reporting outdoor exposures only (aOR=0.68 [0.20-2.22]; **Figure 3**). For participants reporting exposures lasting ≥3 hours, adjusted odds of mask usage were 61% (8-83%) lower among cases than among controls, while for exposures lasting <3 hours, adjusted odds of mask usage were 38% (–34% to 72%) lower among cases than among controls. For exposures without physical contact and those occurring with potentially-infected individuals from outside participants’ households, adjusted odds of mask usage were 59% (7-78%) and 55% (16-76%) lower among cases than among controls, respectively, whereas we did not identify statistically significant evidence of protection from masking in the context of physical encounters or exposures involving household contacts. Among unvaccinated or partially vaccinated participants, adjusted odds of mask usage were 54% (17-77%) lower among cases than controls, whereas among fully vaccinated participants, adjusted odds of mask usage were 22% (–125% to 72%) lower among cases than controls.

**Figure 3.**
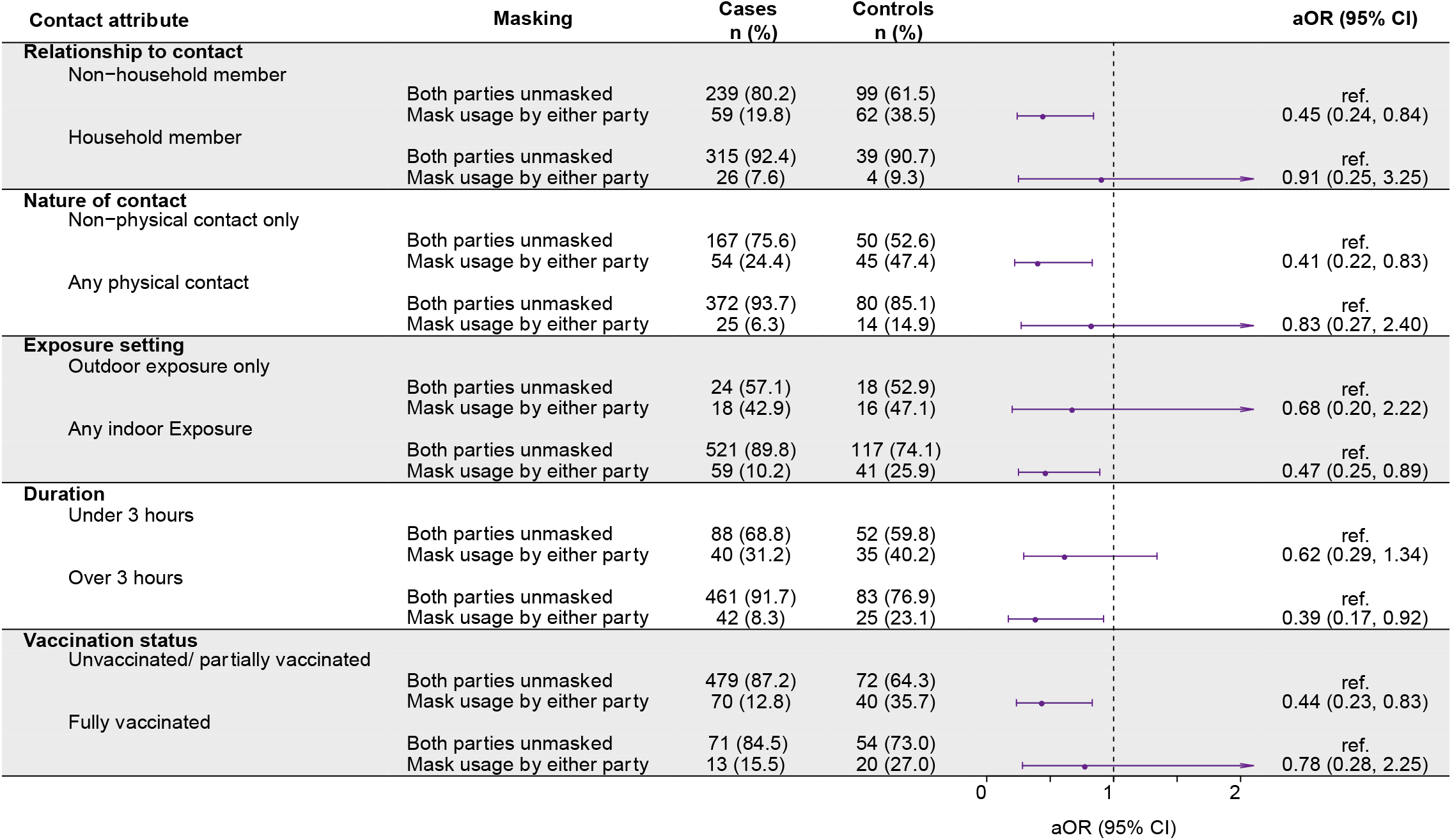
Protective effects of mask-wearing in differing high-risk exposure contexts. aOR: adjusted odds ratio, computed using conditional logistic regression models adjusting for vaccination status of respondent, community exposures (listed in main text), characteristics of the high-risk contact (as listed in **Figure 1**), and participants’ age, sex, and region of residence. An interaction term was included between mask usage and the contact attribute in five separate models. Regression strata were defined for county reopening tiers and week of SARS-CoV-2 test. The aOR represents the adjusted odds ratio for case status comparing mask usage within each category (with respect to relationship, physical/non-physical nature of contact, indoor/outdoor exposure, duration, and participant vaccination status).

Contrary to the hypothesis that masking may reduce individuals’ risk of symptoms, given infection, adjusted odds of mask usage by either participants or their contacts during high-risk exposures were 2.11 (1.01-4.30) fold higher among symptomatic than asymptomatic cases (**Table 4**; **Table S8**). Likewise, point estimates for this association were >1 when disaggregating exposures according to whether masks were worn by participants, their contacts, or both parties. The aOR estimate for mask usage during high-risk interactions was 0.74 (0.36-1.55) for cases who sought care with a medical provider, as compared to cases who did not. Neither the presence of symptoms nor the number of symptoms reported among cases testing positive were associated with high-risk exposures having occurred indoors versus outdoors, having involved household members or other contacts, having involved physical contact or no physical contact, or having lasted ≥3 hours or <3 hours (**Table S9; Table S10**).

**Table 4.** Comparison of infection severity among cases who reported high-risk exposures with and without mask usage.

|  | Symptoms experienced |  |  | Level of care sought |  |  |
| --- | --- | --- | --- | --- | --- | --- |
|  | No symptoms<br>*n* (%)<br>*N*=178 | Symptoms<br>*n* (%)<br>*N*=465 | aOR (95% CI) | No care<br>*n* (%)<br>*N*=499 | Care<br>*n* (%)<br>*N*=144 | aOR (95% CI) |
| Mask usage at interaction |  |  |  |  |  |  |
| No mask usage at interaction | 160 (90.0) | 396 (85.2) | -- | 428 (85.8) | 128 (88.9) | -- |
| Mask usage by either party | 17 (9.6) | 68 (14.6) | 2.11 (1.01, 4.30) | 69 (13.8) | 16 (11.1) | 0.74 (0.36, 1.55) |
| Mask usage by participant and contact |  |  |  |  |  |  |
| Mask usage by both parties | 10 (5.6) | 35 (7.5) | 2.13 (0.83, 5.51) | 37 (7.4) | 8 (5.6) | 0.95 (0.38, 2.42) |
| Mask usage by participant only | 3 (1.7) | 20 (4.3) | 1.73 (0.48, 6.30) | 18 (3.6) | 5 (3.5) | 0.80 (0.22, 2.90) |
| Mask usage by contact only | 4 (2.3) | 13 (2.8) | 2.13 (0.83, 5.51) | 14 (2.8) | 3 (2.1) | 0.36 (0.07, 1.78) |

## DISCUSSION

Our study provides evidence supporting both vaccination and NPIs as strategies to reduce risk of SARS-CoV-2 infection. Among participants who reported recent high-risk exposures, use of face masks was associated with reduced odds of testing positive for SARS-CoV-2 infection. Interacting in an indoor setting, longer (≥3 hour) lengths of interaction, and exposures involving household members were each associated with increased odds of testing positive for SARS-CoV-2 infection among participants who were not fully vaccinated. Excess infection risk associated with each of these exposures was mitigated among fully vaccinated participants. These findings may inform the use of NPIs in populations with limited vaccine access or those ineligible to be vaccinated, and in response to changing epidemiologic conditions such as emergence of variants associated with enhanced infectiousness.

We identified stronger evidence of protection associated with mask usage when contact occurred indoors (vs. outdoors) and when contact lasted ≥3 hours (vs. shorter durations), suggesting mask usage may be of greater relative importance in high-risk circumstances or those where other NPIs cannot be implemented. Whereas mask usage was protective in interactions where participants reported no physical contact with a potentially infectious individual, we did not identify protection in interactions where physical contact was made. Mask usage was also less clearly protective when participants were exposed to a potentially infected member of their own household. This finding may owe to the same factors or reflect the difficulty of adhering to stringent masking over periods of extended or repeated exposure, as may occur among household members [28,29].

Benefits of masking were greatest for unvaccinated participants, among whom we estimated a 56% reduction in odds of infection associated with mask usage during high-risk exposures. We also identified 22% lower odds of infection associated with mask wearing among vaccinated participants. However, analyses within this stratum were underpowered, as most enrollment in this study occurred when vaccine coverage was low and before expansion of the Delta variant, which has been associated with increased transmission risk.

Contrary to prevailing hypotheses [24], symptomatic cases were not more likely than asymptomatic cases to report unmasked, indoor, long-lasting, or physical interactions with their potentially-infected contacts. Limitations of our study included the potential for symptoms reporting to vary among participants, and the possibility that participants may have been pre-symptomatic at the time of their interview/response. In addition, bias may have occurred if individuals’ decision to wear masks was associated with their likelihood of seeking testing when asymptomatic or minimally symptomatic. Direct measurement of SARS-CoV-2 exposure intensity and clinical status was not possible under this design. However, based on our observations, real-world effects of masking and other non-pharmaceutical mitigation measures may have greater impact on individuals’ risk of infection than their likelihood of experiencing symptoms, once infected. Studies in animal models have likewise provided inconsistent support for the hypothesis that reducing SARS-CoV-2 exposure dose may lower the risk of severe disease, given infection [26].

Additional factors which may have modified the likelihood of transmission during high-risk exposure could include the vaccination status of infected contacts [30], the type of masks or face coverings used [31], the physical distance individuals maintained while interacting, and ventilation of indoor spaces where interactions occurred. Obtaining reliable information on these details of each interaction was not feasible through retrospective interviews with participants. While our sample size was under-powered to distinguish between protection associated with masking by participants, their contacts, or both parties, confounding may also arise if the decision to wear masks was influenced by factors we did not measure, including contacts’ vaccination status. This may bias effect size estimates from our study toward the null, along with several other factors including exposure misclassification resulting from our reliance on self-reported behaviors, imperfect knowledge of contacts’ infection status, and the possibility that participants were infected through interactions other than the high-risk exposure events analyzed here.

Our findings provide real-world evidence that NPIs including mask usage reduce risk of SARS-CoV-2 transmission when infectious and susceptible individuals come into contact. We also demonstrate substantial vaccine effectiveness against SARS-CoV-2 in the context of high-risk interactions, suggesting such exposures are not associated with heightened risk of vaccine failure. Study participants were mainly enrolled prior to the Delta variant becoming the predominant SARS-CoV-2 lineage in California. Nonetheless, multiple observational studies have confirmed persistence of vaccine protection against SARS-CoV-2 infection despite the emergence and circulation of new variants [32], and high vaccine effectiveness against severe outcomes including hospitalization and death when post-vaccination infections occur [33]. Amid efforts to increase vaccine uptake as a primary public health strategy, our findings indicate NPIs can protect unvaccinated persons and may also be valuable for vaccinated persons as measures to reduce SARS-CoV-2 transmission.

## Data Availability

Direct data requests to the corresponding author at jlewnard{at}berkeley.edu

## ACKNOWLEDGMENTS

Members of the California COVID-19 Case-Control Study Team include: Helia Samani, Nikolina Walas, Timothy Ho, Erin Xavier, Diana J. Poindexter, Najla Dabbagh, Michelle M. Spinosa, Shrey Saretha, Adrian F. Cornejo, Hyemin Park, Christine Wan, Miriam I. Bermejo, Amanda Lam, Amandeep Kaur, Ashly Dyke, Diana Felipe, Maya Spencer, Savannah Corredor, and Yasmine Abdulrahim.

## Supplementary information

## File S1. Survey Questionnaire

### SECTION 1: INTRODUCTION (∼2 min)

1. **Hello, my name is [**____**] and I am calling on behalf of California Department of Public Health to ask some questions regarding [NAME]’s recent COVID-19 test on [INSERT DATE OF TEST]**.
2. *Make sure you’re on the phone with the correct person*.

*If case is a child under 18y, make sure you are speaking to a parent/ guardian:*

**2a. Am I speaking to [NAME]’s parent or guardian?**

[If yes, proceed to **section 2**]

[If no, proceed to 2b]

**2b. Can you please pass the phone to [NAME]’s parent or guardian?**

[If yes, proceed to **section 2**]

[If no, end call]

*If case is someone older than 18y:*

**2c. Am I speaking to [NAME]?**

[If yes, proceed to **section 2**]

[If no, proceed to 2d]

**2d. Can you please pass the phone to [NAME]?**

[If yes, proceed to **section 2**]

[If no-end call]

*NOTE on proxy respondents:*

> *If an individual is hospitalized or otherwise too sick to answer questions on their own behalf, a caretaker may serve as a proxy respondent, but verbal consent must first be obtained from the primary case both to participate in the study and to have the proxy respondent answer on their behalf*.
>
> *A proxy respondent who speaks English or Spanish may answer if the individual is unable to easily complete the interview in one of these two languages, provided they are able to speak English or Spanish with sufficient proficiency to provide verbal consent for both participation and for communicating via the proxy respondent*.

### SECTION 2: ASSENT (∼1 min)

*If you are speaking to a parent or correct person for the first time, add your name and affiliation before starting:* **Hello, my name is [**_____**]and I am calling on behalf of California Department of Public Health**.

1. **Hi! We are interested in asking you some questions about [YOUR or INSERT CHILD’S NAME] recent COVID-19 test. We are hoping to interview you to try to better understand the spread of COVID-19. Do you have some time to chat?** INTERVIEWER: pause and wait for person to confirm that they are still on the line, check YES if they say they are willing to chat If they do not have time, select NO
2. **So before we start, I want to make sure you understand that everything I ask you is confidential, protected by California’s strict privacy laws, and is only being used to inform public health. Your answers will not be shared with any other federal, state, or local authorities, and you’re welcome to decline to answer any question. We anticipate this will take about 20 minutes. I know that sounds like a long time, but we really appreciate your time and your answers will help us answer some extremely important questions about COVID-19**. **Do you understand the information I have just shared with you?** INTERVIEWER: check “yes” if the respondent answers yes and if you deem the respondent to be competent to proceed with consent and interviewing; check “no” and thank the respondent for their time if the respondent says no or if you deem the respondent is not competent to proceed with consent and interviewing.] If it seems like the person needs a <u>proxy respondent</u> due to not speaking well enough English or being too sick, you may ask “**Is there anyone who can help you answer my questions?”**. If you get the proxy respondent on the phone, re-introduce yourself by starting at the top of Section 2 with “Hello, my name is…” and add at the end, **“Can you help answer questions on [insert name of case/control’s] behalf?”** NOTE that a proxy respondent must be over the age of 14. *Interviewers then seek consent from the participant, but the question asked will depend on the age of the desired case/control*. [If participant is answering on their own behalf AND they are older then 18] **Great, thank you! To confirm, are you willing to participate in this interview?** [If participant is a child older than 14, answering on their own behalf, first ask for consent from the parent for the child to answer the survey] **Great, thank you! I want to let you know that your child [INSERT CHILD’S NAME] may answer questions on their own behalf. Are you willing to allow [INSERT CHILD’S NAME] to participate in this interview? If not, you can answer questions on their behalf**. *Interviewer: if the child older than 14 joins the call, make sure to reintroduce yourself and explain the purpose of the survey*. [If participant is a child younger than 14, and adult is answering on their behalf] **Great, thank you! Are you willing to answer questions about [INSERT CHILD’S NAME]’s recent exposures as part of this interview?** [If a proxy respondent will answer on behalf of the study participant] **If you are able, I would suggest putting the phone on speakerphone during this interview, so [insert name/ relationship of proxy respondent] can help you**. **[INSERT NAME OF CASE-CONTROL], are you willing to participate in this interview?** **[INSERT NAME OF CASE-CONTROL], do you consent to allow [NAME OF PROXY RESPONDENT] to answer my questions during this interview. Please stay close by [NAME OF PROXY RESPONDENT] in case it is necessary to clarify any points that come up**. [If no or asks to be called back later, proceed to end of the survey] [If consent is provided and case/control is 7-18 years old, proceed to 3] *Interviewer: select the following options based off of the consent pattern:*
  - Participant provided consent on their own behalf
  - Parent provided consent for child <18 yrs
  - Participant provided consent for proxy respondent to answer on their own behalf
  - No consent was provided
3. **No problem. But before we hang up, do you mind quickly sharing why you are unable or unwilling to complete this call?** *Record the free response* [**<u>End call</u>**]
4. **[INSERT CHILD’S NAME] is welcome to stand by or join the call to help answer questions**.

[If child joins the call, proceed to 3b, otherwise skip to **section 3**]

**3b. Hi [INSERT CHILD’s NAME]. My name is [<u>______</u>] and I work with the California Department of Public Health. I’m going to ask you some questions about activities in the past couple of weeks. Are you willing to answer these questions so that we can better understand the spread of COVID-19?**

[Proceed to **section 3**]

### SECTION 3: LAST COVID TEST (∼3 min)

1. **Great, so to start, I want to ask whether you know your COVID-19 test result from [INSERT DATE OF TEST]?** *Record whether they know or don’t know their test result by selecting on of the options:*
  - Subject knows test result and is positive
  - Subject knows test result and is negative
  - Subject does NOT know test result and is positive
  - Subject does NOT know test result and is negative
  [If yes and they are positive, proceed to **section 4**] [If yes and they are negative, proceed to 3] [If no, and they are negative, proceed to 2] [If no, and they are positive, proceed to 4]
2. **Your COVID-19 test result from [INSERT DATE OF TEST] has come back negative**. *Record one of the following options:*
  - Yes
  - No
  - Don’t know
  - Refuse
  [Proceed to 3]
3. **Have you ever received a *positive* COVID-19 test result or been told by a health care provider that you are positive for COVID-19?** [If no, proceed to **section 4**] [If yes, end-call saying: **Thanks for letting me know. Those are all the questions I have for you. Thank you for your time and I hope you have a nice day**.
4. **Your COVID-19 test result from [INSERT DATE OF TEST] has come back positive. This means you do have coronavirus disease or COVID-19. In my role with CDPH, I cannot provide you with medical advice. If you need any medical information, please call your healthcare provider. One thing I want to be sure of today is that we have a plan for you to follow up with your healthcare provider, so that they can check on any symptoms you may have and assess your risks. Even if you feel okay now, it is important to have someone you can call if you start feeling sick. If you do not have a healthcare provider, you can go to an urgent care facility or the emergency room if you are not getting better or you feel like you are getting worse**. [Proceed to **section 4**] [If the person brings up clinical questions or concerns about their positive test] **Thank you for sharing that concern. In my role with CDPH, I am not able to give you medical advice. I do want to be sure that you get the help you need. If you believe you are having a medical emergency, you should call 911. Some warning signs that you should go to the emergency room for are: trouble breathing, bluish lips or face, pain or pressure in the chest that does not go away, new confusion or trouble waking or staying awake, but there are other symptoms too. Otherwise, you should call your healthcare provider**.

### SECTION 4: REASONS FOR TESTING (∼3 min)

1. **Next, I’m going to ask you some questions about your COVID-19 test. Can you describe to me why did you choose to get tested on [INSERT DATE OF TEST]?** *Interviewers will select check boxes from the respondent based off of their response, without prompting them from the following list, and will use a write-in option for any additional reasons for seeking testing. After choosing the best answer from the list, confirm your choice the case/control (ex. “So you got tested for pre or post-travel screening?”)*
  - I had contact with someone who tested positive
  - I had contact with someone who had symptoms, but I do not know if they were confirmed to be positive
  - I was told by a public health worker to get tested because I was exposed to a case
  - I was concerned about symptoms I experienced
  - Someone in my household had contact with someone who was positive
  - A person in my household had contact with someone who had symptoms or suspected they had COVID, but we do not know if they are confirmed to be positive.
  - Routine screening for my job
  - Pre or post-travel screening
  - Test required for a medical procedure
  - I just wanted to see if I was infected
  - Don’t know
  - Refuse
  - Other [*interviewer writes in response*]
2. **At the time you were tested on [DATE OF TEST] were you experiencing any COVID-19 symptoms?** *Record Yes/No/Not Sure/ Refuse* [If yes, ask <u>question 4</u>] [If no, proceed to <u>question 5</u>]
3. **Can you please list the symptoms you were experiencing on or 14 days prior to your test on [DATE OF TEST]** *Interviewers will select the symptoms the individuals indicated that they were experiencing. When the respondent is done listing symptoms, the interviewer may prompt, “Are you sure those were all the symptoms you experienced?” and proceed to confirm absence of the 6 most common symptoms (as applicable), in a conversational manner: “No fever, no chills, no muscle pain, no loss of appetite, no shortness of breath, no cough?”* *Select from the following list of symptoms:*
  - Blocked nose
  - Chills
  - Cough
  - Chest pain
  - Diarrhea
  - Muscle pain
  - Fever
  - Headache
  - Hoarseness
  - Loss of appetite
  - Loss of taste
  - Loss of smell
  - Myalgia (muscle pain)
  - Nausea
  - Runny nose
  - Shortness of breath
  - Sneezing
  - Sore throat
  - Stomach pain
  - Sinus pain
  - Sweating
  - Swollen glands
  - Tickle in throat
  - Watery eyes
  - Don’t know
  - Refuse
  - Other
4. **I am now going to read a list of places you may have sought treatment or advice prior to your test on [DATE OF TEST]. After I read the following options, please answer “Yes” or “No”**. *Record Yes/No/ Not Sure/Refuse e for each of the options below*
  - **Did you seek care at an in-person appointment with your usual physician or healthcare provider**
  - **Did you seek care at a telehealth visit or phone appointment with your usual physician or healthcare provider**
  - **Did you seek care at an in-person visit to an urgent care clinic**
  - **Did you seek care at an in-person visit to a healthcare provider at a retail pharmacy**
  - **Did you visit the emergency room?**
  - **Were you admitted to the hospital?**
  - **And just to follow-up, where there any other forms of healthcare from which you sought treatment advice at the time you had your test on [Insert date of test](specify):______________________________**
5. **In the 14 days prior to your test (between ADD DATE to ADD DATE) do you know whether you had known or suspected contact with one or more people who may have tested positive for COVID-19?**

*Select one of the following options*

- Yes-contact with one person who was confirmed positive
- Yes-contact with more than one person who was confirmed positive
- Yes-contact with one person who I suspected was positive
- Yes-contact with more than one person who I suspected was positive
- No known or suspected contact with a positive case
- Not sure
- Refuse

[If case indicated they had KNOWN or SUSPECTED Contact, proceed to **section 5, <u>part A</u>**],

[If the case did not have known or suspected contact, proceed to **section 6**]

### SECTION 5: CONTACT WITH KNOWN OR SUSPECTED CASE (∼8 min)

[If case indicated they had KNOWN or SUSPECTED Contact, proceed to A]

[If case indicated they did NOT have known or suspected contact, proceed to **section 6**]

A. **I’m going to now ask you some questions about the type of contact you had with the person (people) who may have had COVID-19. We are trying to understand sources of exposure and are hopeful that you are willing to answer the questions honestly, knowing that we aren’t looking or expecting any sort of answer**.
  1. **Was the known/ suspected contact someone who lives in your household?** *if plural (contact with >1 person*): **Were any of the known/ suspected contact people who lives in your household** *Record Yes, No, Don’t know, Refuse*
  2. **Did the known/ suspected contact occur indoors, outdoors, or both indoors and outdoors?** *if plural (contact with >1 person):* **Did the known/suspected contacts occur indoors, outdoors, or both indoors and outdoors?** *Record Indoors, Outdoors, Both indoors and outdoors, Unknown, or Refuse*
  3. **In the 14 days prior to your test (between [ADD 14 DAYS – TEST DATE HERE] to [ADD TEST DATE]), what are the locations where you may have had contact with this person?** *if plural:* **In the 14 days prior to your test (between [ADD 14 DAYS – TEST DATE HERE] to [ADD TEST DATE]), what are the locations where you may have had contact with these people)** *Record the free response answer*
  4. **I am now going to ask you about different precautions you may or may not have been able to take when you came into contact with the known or suspected positive case. Please answer “Yes, No or Not Sure” after each question:** *Record Y/ N/ Not sure for each of the options below:*
    - **Did you come within 6 feet of this person, indoors?** *If plural:* **Did you come within 6 feet of any of these people, indoors?**
    - **Did you come within 6 feet of this person, outdoors?** *If plural:* **Did you come within 6 feet of any of these people, outdoors?**
    - **Did you have physical contact with this person, (ie. handshake, hug)?** *If plural*: **Did you have physical contact with any of these people (ie. handshake, hug)**
  5. **Did you wear a mask the entire time, most of the time, some of the time, or none of the time that you interacted with this person?** *If plural*: **Did you wear a mask the entire time, most of the time, some of the time, or none of the time that you interacted with these people?** *Record which of the statements they agree with from below:*
    - I wore a mask the entire time I interacted with this (these) person(s)
    - I wore a mask most of the time I interacted with this (these) person(s)
    - I wore a mask some of the time I interacted with this (these) person(s)
    - I did not wear a mask during this (these) interaction(s)
    - Not sure
    - Refuse
  6. **Did the person you had known or suspected contact with wear a mask the entire time, most of the time, some of the time, or none of the time when you interacted with them?** *If plural:* **Did the people you had known or suspected contact with wear a mask all, most, some, or none of the time that you interacted with them** *Record which of the statements they agree with from below:*
    - They wore a mask the entire time we interacted
    - They wore a mask most of the time we interacted
    - They wore a mask some of the time we interacted
    - They did not wear a mask during this interaction
    - Not sure
    - Refuse
  7. **Did you spend more than 3 consecutive hours with this person in the 14 days prior to your test (between Date to Date)**.
  *If plural*: **Did you spend more than three consecutive hours with these people in the 14 days prior to your test (between Date to Date)** *Record Yes/ No/ Don’t know/ Refuse*

[proceed to **section 6**]

### SECTION 6: EXPOSURE WITH CONTACT KNOWN OR SUSPECTED CASE (∼10 min)

**Next, I want to learn about potential sources of exposure to COVID-19 in the 14 days before your last test: from [ADD 14 DAYS – TEST DATE HERE] to [ADD TEST DATE]. It may help you to pull up a calendar to remember what you were up to over the last two weeks. This chunk usually takes the longest, so thank you in advance for your time**

*Only read the following if they did <u>not</u> have known or suspected contact:*

**[We are trying to understand sources of exposure and are hopeful that you are willing to answer the questions honestly, knowing that we aren’t looking or expecting any sort of answer.]**

1. **I am now going to ask you about a series of locations which you may have visited. After I announce each location, please tell me “Yes, No, or Not sure” to indicate whether you visited that location between [ADD 14 DAYS – TEST DATE HERE] to [ADD TEST DATE]**.
  - **First, did you attend a health appointment or health facility (other than where you got tested for COVID-19)**
  - **Did you go grocery shopping?**
  - **Now I am going to ask you about the times you went to restaurants. Did you go to any restaurants to pick up take-out or to eat at the restaurant?** *Record one of the following options: a) Dine-in (eat at restaurant) only, b) Take-out only, c) Both dining-in and take-out, d) Neither dine-in or take-out, e) Not sure, f) Refuse*
  *If yes and take-out:*
  - **How many times did you get take-out?**
  - **Did you ever have to go inside the restaurant to either place or pick up your take-out order?** *Record one of the following options: a) Yes, I went inside the restaurant either to place or pick-up my order, b) No I did not go inside the restaurant either to place or pick-up my order, d) No I did not go inside the restaurant either to place or pick-up my order, but someone I went to the restaurant with had to go inside to place or pick-up the order, e) not sure, f) refuse*
  *If yes and dine-in:*
  - **How many times did you eat at an <u>indoor</u> restaurant?**
  - **How many times did you eat at an <u>outdoor</u> restaurant?**
  [Skip the following question chunk about bars if respondent is under 21]
  - **Did you attend any bars, breweries, or wine bars?** *If yes, ask:* **Did you attend a bar, brewery or wine bar?** *Select all For each of the places they indicated that they visited:*
  - **How many times did you attend a [bar/brewery/wine bar]?**
  - **When you went to a (those) [bar(s)/brewery(ies)/wine bar(s)], did you spend most of your time indoors, outdoors, or both indoors and outdoors?**
  - **Did you ever visit a coffee shop?** *If yes, ask:*
  - **When you (typically) visited the coffee shop(s), did you have to go inside to place your order?** *Record one of the following options: a) I went inside to place the order, b) I typically placed the order outside or remotely (via. App, web portal, phone order), c) Don’t know, d) refuse*
  - **When you visited a coffee shop, did you (typically) consume your beverage inside the shop, outside the shop, or did you just pick-up the beverage for take-away**. *Record one of the following options a) consumed inside the shop, b) consumed outside the shop (ex. restaurant set up outdoor tables/ chairs and I drank/ate at those tables), c) Got beverage for take-away, d) Don’t know, e) refuse*
  - **Did you go retail shopping?** *If yes, ask:* **And did you go indoor or outdoor retail shopping?**
  - **Did you exercise at gym?** *If yes, ask:* **And was this an indoor or an outdoor gym?**
  - **Did you participate in a group recreational sport (tennis, soccer, basketball, swimming)**
  - **Did you ever leave your house to go for a walk, run, hike or ride a bike outside?** *If yes ask:* **Did you hike, run, walk, or bike with anyone outside your household?** *Select one of the following options a) No, I always hiked, ran, walked, or biked by myself, b) No, but I sometimes/ always ran, walked, or biked with other people who live in my household, c) Yes I hiked ran, walked, or biked with someone who doesn’t live in my household, d) Don’t know, e) Refuse*
  - **Did you ride public transit?**
  - **Did you use a ride share (eg. Taxi, Uber, Lyft, or carpool with individuals who are not members of your household) ?**
  - **Did you fly on a plane?**
  - **Did you attend a parade, rally, march, or protest?**
  - **Did you receive services at a salon or barber?**
  - **Did you attend an indoor movie theater?**
  - **Did you attend a worship service?** *If yes ask:* **And was this an indoor or an outdoor worship service?**
  - **Did you visit or stay at a school, daycare or preschool?** *If yes, ask:* **Was the school or daycare public or private?**
  - **Did you visit a jail, prison, or correctional facility?**
  *If a participant answers yes to any of the questions in 1, follow-up with:* **How many times did you attend [INSERT LOCATION] between [ADD 14 DAYS – TEST DATE HERE] to [ADD TEST DATE]**. **I am now going to ask you (a couple more) some questions about face mask usage between date to date**.
2. **Between [ADD 14 DAYS – TEST DATE HERE], at all of the <u>indoor</u> places we discussed earlier, did you wear a face mask all, most, some, or none of the time?**
  - I wore a face mask all of the time
  - I wore a face mask most of the time
  - I wore a face mask some of the time
  - I never wore a face mask in indoor places
  - I did not go inside any indoor places other than my home
  - I was not in contact with anyone
3. **Between [ADD 14 DAYS – TEST DATE HERE], at all of the <u>indoor</u> places we discussed earlier, did people you came within 6 feet of wear a face mask all, most, some, or none of the time?**
  - They wore a face mask all of the time
  - They wore a face mask most of the time
  - They wore a face mask some of the time
  - They never wore a face mask in indoor places
  - I did not go inside any indoor places other than my home
  - I was not in contact with any people outside my household in indoor places
4. **Between [ADD 14 DAYS – TEST DATE HERE], at all of the <u>outdoor</u> places we discussed earlier, did you wear a face mask all, most, some, or none of the time?**
  - I wore a face mask all of the time
  - I wore a face mask most of the time
  - I wore a face mask some of the time
  - I never wore a face mask in indoor places
  - I did not go inside any outdoor places other than my home
5. **Between [ADD 14 DAYS – TEST DATE HERE], at all of the <u>outdoor</u> places we discussed earlier, did people you came within 6 feet of wear a face mask all, most, some, or none of the time?**
  - They wore a face mask all of the time
  - They wore a face mask most of the time
  - They wore a face mask some of the time
  - They never wore a face mask in indoor places
  - I did not go inside any outdoor places other than my home
  - I was not in contact with any people outside my household in outdoor places
6. **I am now going to ask you some questions about social gatherings. These include any informal gatherings with friends or family who are NOT members of your household). Did you attend any social gatherings between (14 days prior to test result to test result date)?** *Interviewer: note that our definition of social gatherings is mixing with people who don’t otherwise live in your household. If someone had a longer-term family* together *(ie. traveled to visit relatives, but stayed for multiple days, count this as ONE event)*.

> *If yes, ask:* **When you attended social gatherings, were they indoors, outdoors, or both indoors and outdoors?**
>
> *An <u>outdoor only</u> gathering means the person spent the majority of their time outside An <u>indoor only</u> gathering means the person spent the majority of their time*
>
> *A gathering that was “<u>both indoors and outdoors</u>” means the participant was both inside and outside during the social gathering (ex. Sandy had some friends over for dinner and they ate outside on the patio, and then watched a movie in their living room together)*
>
> *If they indicate they attended indoor social gatherings***: How many indoor social gatherings did you attend between (14 days prior to test result to test result date)? About how many people attended these gatherings? Did you eat or drink during any of these (or this) gatherings? When you attended this (these) indoor gathering(s), did you wear a face mask all, most, some or none of the time?**
>
> *Interviewer: note that this question about mask usage is distinct from the question earlier*.
>
> *If they indicate they attended outdoor social gatherings:* **How many outdoor social gatherings did you attend between (14 days prior to test result to test result date)? About how many people attended these gatherings? Did you eat or drink during any of these (or this) gatherings? When you attended this (these) outdoor gathering(s), did you wear a face mask all, most, some or none of the time?**
>
> *If they indicate they attended social gatherings that were both inside and outside:* **How many social gatherings did you attend between (14 days prior to test result to test result date) that were both indoor and outdoor? About how many people attended these gatherings? Did you eat or drink during any of these (or this) gatherings? When you attended this (these) outdoor gathering(s), did you wear a face mask all, most, some or none of the time?**

- **Did you attend any other kind of event where there are <u>5 or more</u> people who are not in your household in attendance?** *Interviewer: If necessary, prompt with options like a sporting event, concert, festival, etc*. **Specify the event:______________**

[Proceed to **section 7**]

### SECTION 7: OCCUPATION (∼1 min)

1. **I am now going to ask you some questions about your occupation. Between [ADD 14 DAYS – TEST DATE HERE] to [ADD TEST DATE] did you attend work, school, or volunteering commitments exclusively at home, both at home and in “in-person”, or exclusively “in-person”**.
  - I work, study, and/or volunteer at home
  - I attend work, school, and/or volunteering “in-person”
  - I attend work, school, and/or volunteering both “in-person” and at home
  - I am not currently working, in school, or in a volunteer position.
  [If respondent is a student, skip question and just record “student”]
2. **Can you tell me what your job is?** (*Record open ended response]* [If they attend work, school, or volunteering commitments in person or both at home & in person, proceed to question 3, otherwise proceed to **Section 8**] **3a. Do you come into close contact (within 6 feet) of more than 10 people per day at work/school/volunteering?** *Record: Yes or No* **3b. Do you primarily attend work/school/volunteering indoors, outdoor, or both indoors and outdoors?** *Record: indoors, outdoors, or both*

[Proceed to **section 8**]

### SECTION 8: VACCINATION (∼2 min)

**I am now going to ask you some questions about the COVID-19 vaccine**.

1. **Do you have any conditions that might place you higher risk for COVID-19?** *Interviewers may prompt with examples such diabetes, high blood pressure, overweight, being immunocompromised if requested. Select options from list below*
  - Lung conditions: COPD, lung cancer, cystic fibrosis, moderate to severe asthma, pulmonary fibrosis
  - Heart disease
  - High blood pressure
  - Obesity
  - Overweight
  - Diabetes
  - Weakened immune system: organ transplant, cancer treatment, bone marrow transplant, HIV/AIDS, sickle cell anemia, thalassemia
  - Chronic kidney disease
  - Chronic liver disease
  - Pregnant (first, second, or third trimester)
2. **Have you received any doses of a COVID-19 vaccine?** *Record: Yes, No* [If they have not received any doses, skip to 2b, otherwise ask question 3] **2b. Do you plan to receive any doses of the COVID-19 vaccine?** *Record: Yes, No, not sure, refuse* [If they are not planning to receive any doses or are not sure yet, ask 2c, otherwise, ask skip to **section 9**] **2c. Can you describe to me why you are not planning to receive the COVID-19 vaccine?** *Record reason in check box*
3. **How many doses of the COVID-19 vaccine have you received?** *Record: 1, 2*
4. **Do you have a vaccine card on hand from when you got the COVID-19 vaccine?** *If yes, ask them to get their vaccine card. If no, ask them to do their best remembering and try pulling up a calendar to help them remember*.
5. **What dates did you receive your dose(s)?** *Record the date of each vaccine*
6. **Do you know what product COVID-19 vaccine you received?** *Record the product of each dose*
7. **Do you have access to a COVID-19 vaccination clinic at your work or school?** *Record: yes/ no/ not sure/ refuse*
8. **Where did you get your COVID-19 vaccine?** *Record: mass vaccination site, hospital, nursing home, at my work, at my school, at a retail pharmacy, at a retail shop (eg. Walmart)*
9. **At the time you received the vaccine was it required to attend work or school?** *Record: yes/ no/ not sure/ refuse*

[Proceed to **section 9]**

### SECTION 9: DEMOGRAPHICS (∼5 min)

**I just have a few more questions. Again, anything you share with me is confidential and protected by California’s strict privacy laws. The information we collect about you will assist the health department in their COVID-19 response**.

1. **First, I’m going to ask you some general questions about COVID-19. From the beginning of the pandemic to the time you were tested on [DATE OF TEST], how worried did you feel about getting COVID-19? Would you say you felt:**
  - **Very worried**
  - **Somewhat worried**
  - **Neutral**
  - **Not worried at all**
2. **Since the beginning of the pandemic, there have been a lot of recommendations on behaviors that can reduce the risk of COVID-19 including avoiding large crowds, travel, and maintaining 6 feet of distance in public places. Would you say that you strongly agree, agree, are neutral, disagree, or strongly disagree that these measures reduce the risk of COVID-19?** *Record strongly agree, agree, neutral, disagree, strongly disagree*
3. **Another recommendation to reduce the spread of COVID-19 is wearing face masks. Would you say that you strongly agree, agree, are neutral, disagree, or strongly disagree that face masks reduce the risk of COVID-19?** *Record strongly agree, agree, neutral, disagree, strongly disagree*

**Last, I want to capture some information about demographics**.

1. **So do you mind sharing how old you are?** *Record free response*
2. **Next, please let me know which of the following race/ethnicities best describe yourself. You may select all that apply:**
  - **White**
  - **Black**
  - **Hispanic**
  - **Asian**
  - **Native American or Alaska Native**
  - **Native Hawaiian or other Pacific Islander**
3. **What is your sex/ gender?** *Record Man, Woman, Non=-binary, Prefer to self-describe, Refuse, Don’t know*
4. **What is your zip code of your home address?** *Record address using encryption tool*
5. **What is your home address?** *Record address using encryption tool after verifying it is an address using google maps*
6. **Which of the following best describes your living arrangement:**
  - **Private home**
  - **Apartment, or condominium**
  - **Skilled nursing facility**
  - **College or university student housing**
  - **Military quarters**
  - **Emergency or transitional shelter**
  - **Other (please describe)**
7. **How many people live in your household?**
8. **How many bedrooms do you have in your household?**
9. **Do you have any children under 18 at your home?**
10. **Are any of your children under 18 attending in-person instruction, school, or daycare?**
11. **Does anyone visit your home on a regular basis like a cleaning service or babysitter?** If you are talking to a child aged 14-17, at this point you can end the interview with the child and ask to speak with their parent/guardian. When you get back on the phone with the parent or guardian, you can say something like **[“Hi again, thank you so much for letting me speak with your child, it was extremely helpful. We are wrapping up the survey with some demographic questions and my last question that I didn’t want your child to have to answer was whether you are willing to share your total household income?”]**
12. **What is your total household income? Answer on behalf of everyone you share finances with**. [If you are speaking with POSITIVE case, proceed to 13] [If you are speaking with NEGATIVE control, proceed to 14]
13. **Thank you for participating in this survey. You may be contacted by another staff member at the health department to check in on you. They will ask you questions about your health and well-being to make sure you’re ok**.
14. **Thank you for participating in our survey. We appreciate your time**.

**Table S1:** Counties included in each geographic region.

| County | Region |
| --- | --- |
| Alameda County | San Francisco San Francisco Bay Area |
| Alpine County | Sierras Region |
| Amador County | Sierras Region |
| Butte County | Northern Sacramento Valley |
| Calaveras County | Sierras Region |
| Colusa County | Northern Sacramento Valley |
| Contra Costa County | San Francisco Bay Area |
| Del Norte County | Northwestern California |
| El Dorado County | Sierras Region |
| Fresno County | San Joaquin Valley |
| Glenn County | Northern Sacramento Valley |
| Humboldt County | Northwestern California |
| Imperial County | San Diego and southern border |
| Inyo County | Sierras Region |
| Kern County | San Joaquin Valley |
| Kings County | San Joaquin Valley |
| Lake County | Northwestern California |
| Lassen County | Sierras Region |
| Los Angeles County | Greater Los Angeles area |
| Madera County | San Joaquin Valley |
| Marin County | San Francisco Bay Area |
| Mariposa County | Sierras Region |
| Mendocino County | Northwestern California |
| Merced County | San Joaquin Valley |
| Modoc County | Sierras Region |
| Mono County | Sierras Region |
| Monterey County | Central Coast |
| Napa County | San Francisco Bay Area |
| Nevada County | Sierras Region |
| Orange County | Greater Los Angeles area |
| Placer County | Sierras Region |
| Plumas County | Sierras Region |
| Riverside County | Greater Los Angeles area |
| Sacramento County | Central Valley |
| San Benito County | San Francisco Bay Area |
| San Bernardino County | Greater Los Angeles area |
| San Diego County | San Diego and southern border |
| San Francisco County | San Francisco Bay Area |
| San Joaquin County | San Joaquin Valley |
| San Luis Obispo County | Central Coast |
| San Mateo County | San Francisco Bay Area |
| Santa Barbara County | Central Coast |
| Santa Clara County | San Francisco Bay Area |
| Santa Cruz County | San Francisco Bay Area |
| Shasta County | Northwestern California |
| Sierra County | Sierras Region |
| Siskiyou County | Northwestern California |
| Solano County | San Francisco Bay Area |
| Sonoma County | San Francisco Bay Area |
| Stanislaus County | San Joaquin Valley |
| Sutter County | Northern Sacramento Valley |
| Tehama County | Northern Sacramento Valley |
| Trinity County | Northwestern California |
| Tulare County | San Joaquin Valley |
| Tuolumne County | Sierras Region |
| Ventura County | Greater Los Angeles area |
| Yolo County | Northern Sacramento Valley |
| Yuba County | Northern Sacramento Valley |

**Table S2:** Demographic attributes of study population, stratified by high-risk contact.

|  |  | Total | No High-risk contact |  | High-risk contact |  |
| --- | --- | --- | --- | --- | --- | --- |
|  |  | *n* (%)<br>*N*=2541 | Case<br>*n* (%)<br>*N*=617 | Control<br>*n* (%)<br>*N*=1046 | Case<br>*n* (%)<br>*N*=643 | Control<br>*n* (%)<br>*N*=204 |
| Age | 0-6 | 70 (2.8) | 6 (1.0) | 34 (3.3) | 28 (4.4) | 2 (1.0) |
|  | 7-12 | 102 (4.0) | 14 (2.3) | 40 (3.8) | 37 (5.8) | 11 (5.4) |
|  | 13-17 | 128 (5.0) | 21 (3.4) | 50 (4.8) | 41 (6.4) | 12 (5.9) |
|  | 18-29 | 817 (32.2) | 200 (32.4) | 342 (32.7) | 206 (32.0) | 63 (30.9) |
|  | 30-49 | 887 (34.9) | 231 (37.4) | 353 (33.7) | 202 (31.4) | 89 (43.6) |
|  | 50-64 | 386 (15.2) | 106 (17.2) | 162 (15.5) | 91 (14.2) | 20 (9.8) |
|  | 65+ | 151 (5.9) | 39 (6.3) | 65 (6.2) | 38 (5.9) | 7 (3.4) |
| Sex | Male | 1232 (48.5) | 312 (50.6) | 516 (49.3) | 295 (45.9) | 93 (45.6) |
|  | Female | 1309 (51.5) | 305 (49.4) | 530 (50.7) | 348 (54.1) | 111 (54.4) |
| Household income | Under \$50,000 | 622 (24.5) | 179 (29.0) | 226 (21.6) | 159 (24.7) | 47 (23.0) |
| | \$50,000 to \$100,000 | 563 (22.2) | 134 (21.7) | 219 (20.9) | 160 (24.9) | 41 (20.1) |
| | \$100,000 to \$150,000 | 304 (12.0) | 50 (8.1) | 152 (14.5) | 63 (9.8) | 36 (17.6) |
| | Over \$150,000 | 345 (13.6) | 72 (11.7) | 167 (16.0) | 73 (11.4) | 30 (14.7) |
|  | Refuse | 434 (17.1) | 113 (18.3) | 171 (16.3) | 114 (17.7) | 32 (15.7) |
|  | Not sure | 273 (10.7) | 69 (11.2) | 111 (10.6) | 74 (11.5) | 18 (8.8) |
| Race/ ethnicity | Non-Hispanic White | 1061 (43.1) | 249 (41.4) | 440 (43.2) | 272 (44.2) | 85 (43.1) |
|  | Non-Hispanic Black | 118 (4.8) | 44 (7.3) | 35 (3.4) | 32 (5.2) | 6 (3.0) |
|  | Hispanic (any race) | 746 (30.3) | 210 (34.9) | 298 (29.3) | 176 (28.6) | 54 (27.4) |
|  | Asian | 237 (9.6) | 51 (8.5) | 116 (11.4) | 51 (8.3) | 16 (8.1) |
|  | Native American | 30 (1.2) | 6 (1.0) | 6 (0.6) | 15 (2.4) | 2 (1.0) |
|  | Native Hawaiian | 17 (0.7) | 2 (0.3) | 10 (1.0) | 5 (0.8) | 0 (0.0) |
|  | More than 1 race | 252 (10.2) | 39 (6.5) | 113 (11.1) | 65 (10.6) | 34 (17.3) |
| Region of residence<sup>1</sup> | *Predominantly urban regions* |  |  |  |  |  |
|  | San Francisco Bay Area | 296 (11.6) | 75 (12.2) | 131 (12.5) | 73 (11.4) | 14 (6.9) |
|  | Greater Los Angeles Area | 283 (11.1) | 76 (12.3) | 118 (11.3) | 70 (10.9) | 19 (9.3) |
|  | Greater Sacramento Area | 285 (11.2) | 55 (8.9) | 117 (11.2) | 82 (12.8) | 25 (12.3) |
|  | San Diego and southern Border | 277 (10.9) | 76 (12.3) | 120 (11.5) | 62 (9.6) | 18 (8.8) |
|  | *Predominantly rural regions* |  |  |  |  |  |
|  | Central Coast | 304 (12.0) | 74 (12.0) | 113 (10.8) | 79 (12.3) | 34 (16.7) |
|  | Northern Sacramento Valley | 277 (10.9) | 66 (10.7) | 119 (11.4) | 69 (10.7) | 19 (9.3) |
|  | San Joaquin Valley | 281 (11.1) | 73 (11.8) | 110 (10.5) | 65 (10.1) | 27 (13.2) |
|  | Northwestern California | 272 (10.7) | 68 (11.0) | 106 (10.1) | 68 (10.6) | 28 (13.7) |
|  | Sierras Region | 266 (10.5) | 54 (8.8) | 112 (10.7) | 75 (11.7) | 20 (9.8) |
| Vaccination status<sup>2</sup> | Unvaccinated | 1617 (66.7) | 483 (80.2) | 521 (53.7) | 502 (79.1) | 89 (47.8) |
|  | Partially vaccinated | 257 (10.6) | 43 (7.1) | 139 (14.3) | 49 (7.7) | 23 (12.4) |
|  | Fully vaccinated | 550 (22.7) | 76 (12.6) | 310 (32.0) | 84 (13.2) | 74 (39.8) |
| County reopening tier<sup>3</sup> | Purple tier (most restrictive) | 599 (23.6) | 141 (22.9) | 245 (23.4) | 160 (24.9) | 41 (20.1) |
|  | Red tier | 649 (25.5) | 164 (26.6) | 270 (25.8) | 163 (25.3) | 40 (19.6) |
|  | Orange tier | 625 (24.6) | 136 (22.0) | 285 (27.2) | 165 (25.7) | 35 (17.2) |
|  | Yellow tier (least restrictive) | 75 (3.0) | 20 (3.2) | 32 (3.1) | 17 (2.6) | 6 (2.9) |
|  | After June 15<sup>th</sup> | 593 (23.3) | 156 (25.3) | 214 (20.5) | 138 (21.5) | 82 (40.2) |
| Symptoms experienced | No symptoms | 1330 (52.3) | 111 (18.0) | 869 (83.1) | 178 (27.7) | 158 (77.5) |
|  | At least one symptom | 1211 (47.7) | 506 (82.0) | 177 (16.9) | 465 (72.3) | 46 (22.5) |

Recent high-risk exposure is defined as reported contact with an individual known or suspected to have been infected with SARS-CoV-2 at any time within the 14 days before participants were tested.
<sup>1</sup>We list counties grouped into each region in **Table S1**.
<sup>2</sup>An individual was considered partially-vaccinated if their SARS-CoV-2 test date with less than 14 days before their second dose of a mRNA vaccine product (Pfizer/BioNTech [BNT-162b2] or Moderna [mRNA-1273]), or less than 14 days after their first dose of a single dose vaccine product (Jansen Pharmaceutical Companies [JNJ-78436735]). An individual was considered fully-vaccinated if their SARS-CoV-2 test date was more than 14 days after their second dose of a mRNA vaccine product (Pfizer/BioNTech [BNT-162b2] or Moderna [mRNA-1273]), or more than 14 days after their first dose of a single dose product (Jansen Pharmaceutical Companies [JNJ-78436735]).
<sup>3</sup>The State of California implemented a tiered system of reopening to reduce risk of SARS-CoV-2 transmission in community settings. On June 15, 2021, California discontinued the tiered system, relaxed facial requirements in certain indoor settings, and allowed businesses to reopen without physical distancing restrictions.

**Table S3:** Location(s) of confirmed or suspected contact among interviewed participants.

| Location of contact | Total participants<br>*n* (%)<br>*N*=847 | Confirmed contact<br>*n* (%)<br>*N*=694 | Suspected contact<br>*n* (%)<br>*N*=153 |
| --- | --- | --- | --- |
| Residence<sup>a</sup> | 456 (53.8) | 383 (55.2) | 73 (47.7) |
| Same household | 332 (72.8) | 282 (73.6) | 50 (68.5) |
| Outside of the household | 80 (17.5) | 64 (16.7) | 16 (21.9) |
| Visits to multiple households | 38 (8.3) | 32 (8.4) | 6 (8.2) |
| Unknown but related to a residence | 6 (1.3) | 5 (1.3) | 1 (1.4) |
| Workplace | 115 (13.6) | 88 (12.7) | 27 (17.6) |
| Public space or event<sup>b</sup> | 96 (11.3) | 78 (11.2) | 18 (11.8) |
| In a vehicle, including public transportation | 14 (1.7) | 12 (1.7) | 2 (1.3) |
| Multiple location types<sup>c</sup> | 112 (13.2) | 94 (13.5) | 18 (11.8) |
| Unknown<sup>d</sup> | 54 (6.4) | 39 (5.6) | 15 (9.8) |

<sup>a</sup> Includes individuals who reported having contact with known or suspected case(s) from the same household, outside of the household, and during visits to multiple households (e.g., participant had contact with known cases from two separate residences)
<sup>b</sup> Excludes individuals who reported working at a public space (e.g., bar, restaurant, hospital) at the time of contact
<sup>c</sup> Includes individuals who reported having contact at any combination of the above location types (e.g., participant had contact with suspected cases at their workplace and at a relative's residence)
<sup>d</sup> Includes individuals who declined to answer or did not report a specific location

**Table S4:** Reasons for testing among participants who reported high-risk interactions.

| Reasons* | Controls |  | Cases |  |
| --- | --- | --- | --- | --- |
|  | Unvaccinated<br>N=89 | Vaccinated<sup>†</sup><br>N=97 | Unvaccinated<br>N=502 | Vaccinated<br>N=133 |
| Contact with positive case | 52 (58.4) | 64 (66.0) | 355 (70.7) | 97 (72.9) |
| Contact with symptomatic individual, unknown whether confirmed positive | 11 (12.4) | 11 (11.3) | 39 (7.8) | 7 (5.3) |
| Told by public health worker to get tested | 0 (0.0) | 4 (4.1) | 5 (1.0) | 2 (1.5) |
| Routine screening | 22 (24.7) | 12 (12.4) | 14 (2.8) | 6 (4.5) |
| Test required for medical procedure or hospital admittance | 2 (2.2) | 0 (0.0) | 5 (1.0) | 0 (0.0) |
| Someone in household had contact with a positive case | 2 (2.2) | 2 (2.1) | 27 (5.4) | 6 (4.5) |
| I just wanted to see if I was infected | 8 (9.0) | 6 (6.2) | 18 (3.6) | 4 (3.0) |
| Concerned about symptoms | 10 (11.2) | 9 (9.3) | 193 (38.4) | 62 (46.6) |
| Pre or post-travel screening | 1 (1.1) | 6 (6.2) | 4 (0.8) | 4 (3.0) |

\*Since interviewers indicated all reasons listed by participants, and some participants refused to respond to the question, reasons will not sum to the total sample size.
<sup>†</sup>An individual is considered vaccinated if they have had at least one dose of any SARS-CoV-2 vaccine product.

**Table S5:** Regression parameter estimates.

| Covariate | aOR (95% CI) |
| --- | --- |
| Restaurant | 0.79 (0.37, 1.71) |
| Bar/ brewery/ winery | 0.79 (0.37, 1.71) |
| Coffee shop | 0.92 (0.55, 1.60) |
| Retail shop | 0.87 (0.53, 1.44) |
| Gym | 0.89 (0.44, 1.82) |
| Ride share service | 0.97 (0.48, 2.03) |
| Public transport | 1.06 (0.32, 3.46) |
| Salon | 0.64 (0.33, 1.26) |
| Worship service | 2.68 (0.76, 9.19) |
| Social gathering | 1.15 (0.73, 1.82) |

**Table S6:**
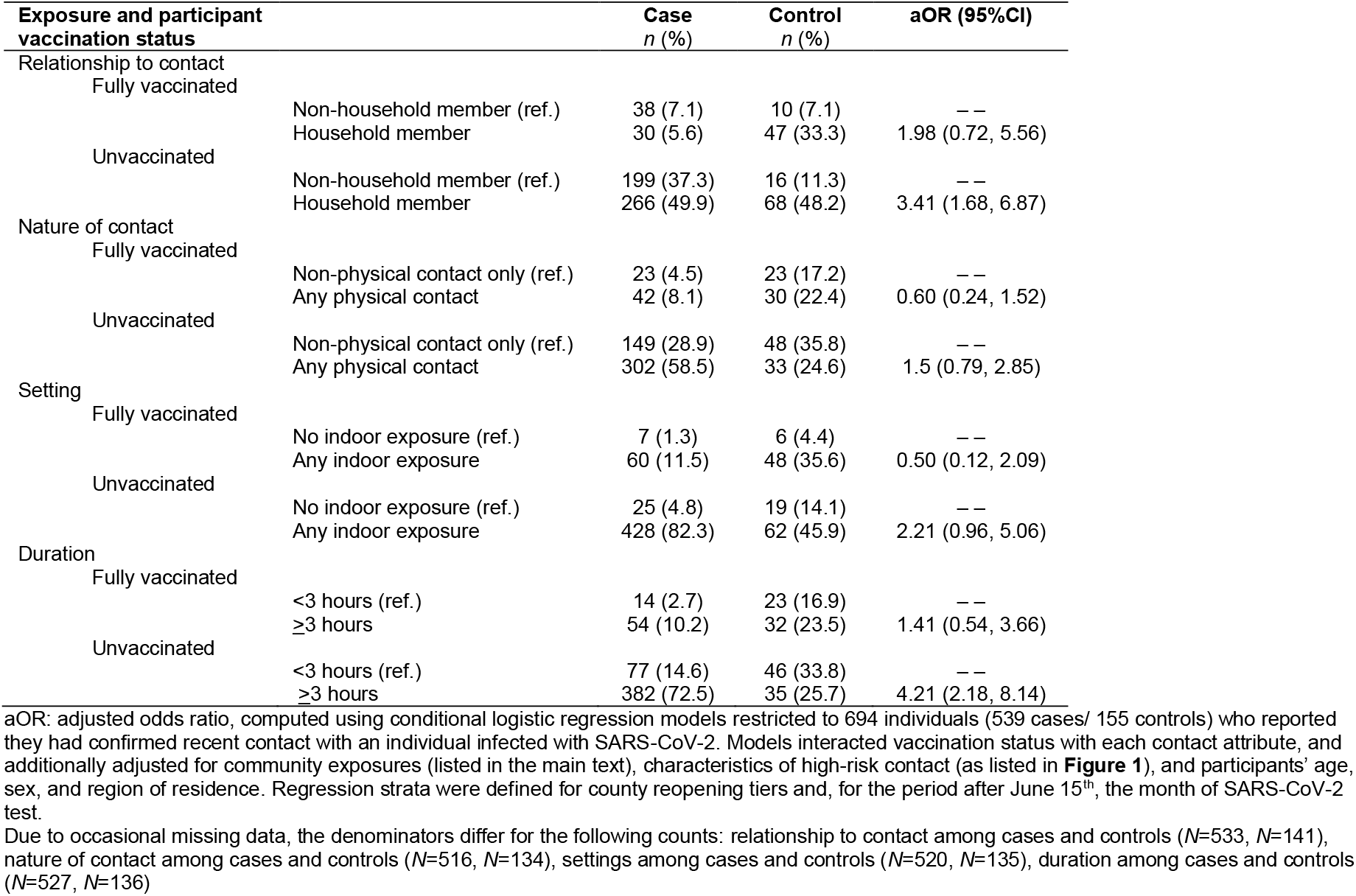
Predictors of infection following high-risk exposure among participants with confirmed SARS-CoV-2 contact.

**Table S7:** Protective effects of mask-wearing and vaccination in the context of high-risk exposure among participants with confirmed SARS-CoV-2 contact.

| Exposure and participant vaccinations status | Case<br>*n* (%)<br>*N*=539 | Control<br>*n* (%)<br>*N*=155 | aOR (95% CI) |
| --- | --- | --- | --- |
| Mask usage at interaction |  |  |  |
| No mask usage at interaction | 473 (87.9) | 103 (66.5) | ref. |
| Mask usage by either party | 65 (12.1) | 52 (33.5) | 0.48 (0.24, 0.95) |
| Mask usage by participant and contact |  |  |  |
| Mask usage by both parties | 473 (87.9) | 103 (66.5) | 0.63 (0.28, 1.47) |
| Mask usage by participant only | 65 (12.1) | 52 (33.5) | 0.13 (0.03, 0.63) |
| Mask usage by contact only | 473 (87.9) | 103 (66.5) | 0.47 (0.15, 1.48) |
| Vaccination status of respondent |  |  |  |
| No vaccine doses | 424 (79.4) | 66 (46.8) | ref. |
| Incomplete vaccination series | 42 (7.9) | 18 (12.8) | 0.38 (0.15, 0.92) |
| Full vaccination series<sup>1</sup> | 68 (12.7) | 57 (40.4) | 0.20 (0.09, 0.41) |

Due to occasional missing data, the denominators differ for the following counts: mask usage at interaction among cases (*N*=538), vaccination status among cases and controls (*N*=534, *N*=141).

**Table S8:** Number of reported symptoms among cases by level of mask usage during high-risk exposures.

| **Mask usage** | **Mean number of reported symptoms (SD)** |
| --- | --- |
| Mask usage at interaction |  |
| No mask usage at interaction | 2.44 (2.05) |
| Mask usage by either party | 2.26 (2.21) |
| Mask usage by participant and contact |  |
| Mask usage by both parties | 2.58 (2.41) |
| Mask usage by participant only | 2.61 (1.64) |
| Mask usage by contact only | 1.82 (1.38) |

Among cases who reported a high-risk interaction, we computed the mean number of reported symptoms within strata of reported mask usage by the participant and their contact during the interaction

**Table S9:**
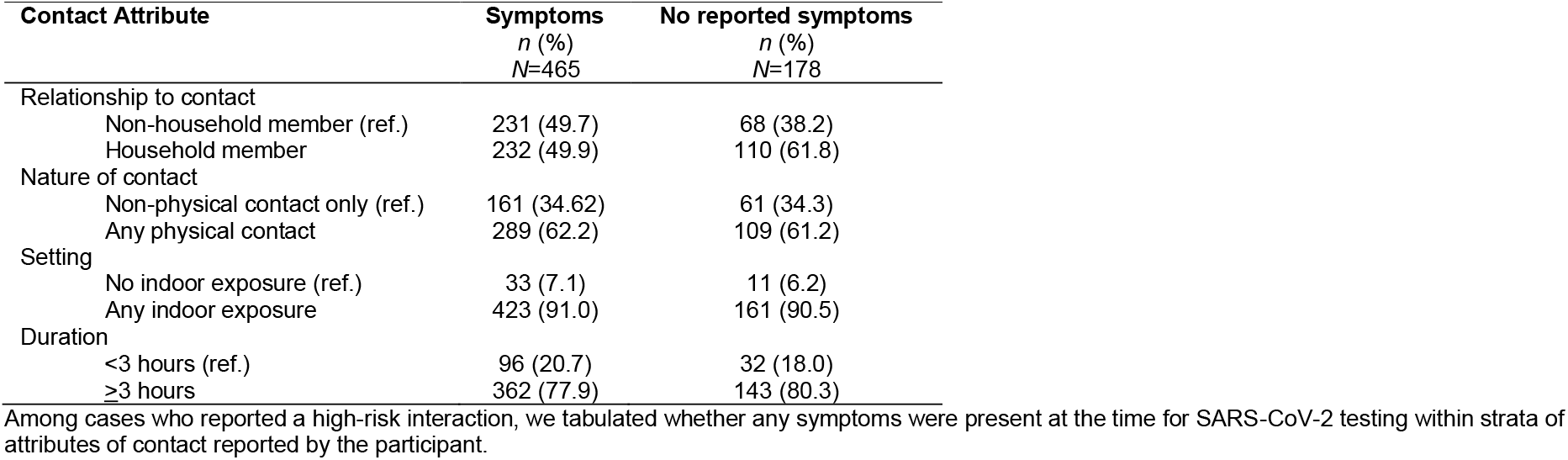
Presence of reported symptoms by attributes of high-risk contact among cases.

**Table S10:** Number of reported symptoms among cases by attributes of the high-risk contact.

| Contact Attribute | Mean number of reported symptoms (SD) |
| --- | --- |
| Relationship to contact |  |
| Non-household member (ref.) | 2.59 (2.22) |
| Household member | 2.01 (2.14) |
| Nature of contact |  |
| Non-physical contact only (ref.) | 2.24 (2.24) |
| Any physical contact | 2.33 (2.19) |
| Setting |  |
| No indoor exposure (ref.) | 2.41 (2.20) |
| Any indoor exposure | 2.29 (2.20) |
| Duration |  |
| <3 hours (ref.) | 2.51 (2.49) |
| ≥3 hours | 2.23 (2.12) |

Among cases who reported a high-risk interaction, we calculated the mean number of symptoms at the time of SARS-CoV-2 testing within strata of attributes of contact reported by the participant.

